# Reaching health facilities in situations of emergency: Experiences of pregnant women in Africa’s largest megacity

**DOI:** 10.1101/2020.03.18.20036830

**Authors:** Aduragbemi Banke-Thomas, Mobolanle Balogun, Ololade Wright, Babatunde Ajayi, Ibukun-Oluwa Abejirinde, Abimbola Olaniran, Rokibat Olabisi Giwa-Ayedun, Bilikisu Odusanya, Bosede Bukola Afolabi

## Abstract

Travel of pregnant women requiring emergency obstetric care (EmOC) to health facilities remains a ‘black box’ of many unknowns to the health system, more so in megacities which are fraught with wide inequalities. This in-depth study on travel of pregnant women in Africa’s largest megacity is based on interviews conducted between September 2019 and January 2020 with 47 women and 11 of their relatives who presented at comprehensive EmOC facilities in situations of emergency, requiring some EmOC services. Despite recognising danger signs, pregnant women are often faced with conundrums on “when”, “where” and “how” to reach EmOC facilities. While the decision-making process is a shared activity amongst all women, the available choice-options vary depending on socio-economic status. Women preferred to travel to facilities deemed to have “nicer” health workers, even if these were farther from home. Reported travel time ranged from 5-240 minutes in daytime and 5-40 minutes at night. Many women reported facing remarkably similar travel experiences, with varied challenges faced in the daytime (traffic congestion) compared to night-time (security concerns and scarcity of public transportation). This was irrespective of their age, socio-economic background, or obstetric history. However, the extent to which this experience impacted on their ability to reach facilities depended on their agency and support systems. Travel experience was better if they had their personal vehicle for travel at night, support of relatives or direct/indirect connections with senior health workers at comprehensive EmOC facilities. Referral barriers between facilities further prolonged delays and increased cost of travel for many women. If the goal to leave no one behind remains a priority, in addition to other health systems strengthening interventions, referral systems need to be improved, advocacy on policies to encourage women to utilise nearby functional facilities when in situations of emergency and private sector partnerships should be explored.

## 1. Introduction

Despite a 29% reduction in global maternal deaths from 1990 to 2015, maternal mortality remains a global health challenge, with about 280,000 women still dying every year due to complications of pregnancy and childbirth. Ninety-nine percent of these deaths occur in low- and middle-income countries (LMICs), with sub-Saharan Africa accounting for 66% of all maternal deaths (GBD 2015 Maternal Mortality Collaborators., 2016; World Health Organization et al., 2015). Evidence in the literature suggests that timely and quality emergency obstetric care (EmOC) provided by skilled health personnel reduces institutional maternal mortality by 15-50% and intra-partum stillbirths by 45-75% (Paxton et al., 2005; World Health Organization et al., 2018).

As described by the World Health Organization (WHO), EmOC is a package of clinical and surgical evidence-based interventions that are most effective in managing the five major causes of maternal deaths, namely, hypertensive disorders, severe bleeding, sepsis, obstructed labour and unsafe abortion (World Health Organization et al., 2009). Seven interventions comprising the administration of parenteral antibiotics, uterotonic drugs, parenteral anticonvulsants, manual removal of placenta, removal of retained products of conception, assisted vaginal delivery and neonatal resuscitation are classified as basic emergency obstetric care (BEmOC). In addition to these BEmOC services, blood transfusions and caesarean sections make up comprehensive EmOC (CEmOC) (World Health Organization et al., 2009).

However, delay in decision-making of women to seek care (first delay), delay in travel of women to health facilities that have capacity to provide the needed care (second delay), and delay in receiving appropriate care upon arrival at the facility (third delay) impede access to EmOC services and have long been associated with increased risk for maternal deaths (Thaddeus and Maine, 1994). Existing evidence suggests that in LMICs, in addition to the direct effect that distance between places of residence and EmOC facilities have on the “second delay” (i.e. increasing the time between decision to access care and its receipt), distance also has an indirect effect on the “first delay”, as it sometimes create a disincentive to seek care (Gabrysch and Campbell, 2009; Lohela et al., 2012; Panciera et al., 2016). Factors relating to actual travel to health facilities also influences women’s choice to use skilled health personnel vs. traditional birth attendants (Vieira et al., 2012).

Several actions can be taken at a health system level to minimise the delays. Skilled health personnel can leverage ante-natal care (ANC) attendance or community outreaches to encourage women to seek facility-based delivery when in an emergency (Baatiema et al., 2019; Devasenapathy et al., 2017; Fekadu et al., 2018; Hill et al., 2019; Sialubanje et al., 2017), thereby forestalling the first delay. In addition, they can provide the care that the women require on arrival at health facilities in a timely fashion (Bailey et al., 2006; Paxton et al., 2005), which reduces the third delay. However, the travel trajectory between home and facility is a ‘black box’ in many LMIC health systems, as women are on their own or with their relatives, and expected to find their way to health facilities in emergency situations, placing them in even more precarious situations (Afari et al., 2014; Hezelgrave et al., 2011).

Indeed, in emergency situations, the path of travel that women take to reach health facilities may be convoluted (Banke-Thomas et al., 2019), which in itself can prolong their travel trajectory (Echoka et al., 2013). This travel path can be even more complicated in megacities, defined as a metropolitan area with a population of more than 10 million people (United Nations, 2019). Megacities are known to be fraught with increasing socio-economic vulnerability due to mounting poverty, socio-spatial, political and institutional fragmentation and often extreme forms of segregation, disparities, and violence (Jowell et al., 2017; Kraas, 2007). But even within mega-cities, wide inequalities warrant a need for further categorisation into slums and non-slum areas (Gaur et al., 2013). In addition, varying geographical terrains, including land and water, pose different travel challenges and accessibility barriers to inhabitants (Kraas, 2007). These considerations point to the non-homogenous nature of megacities and with their rapid emergence, the importance of the second delay is more critical than ever.

In sub-Saharan Africa, where the burden of maternal deaths is highest, two cities - Kinshasa, Democratic Republic of Congo and Lagos, Nigeria have emerged as megacities with population estimates of 12 and 21 million respectively (United Nations, 2019). However, according to a 2018 systematic review that looked at access and utilisation of EmOC at health facilities in sub-Saharan Africa (Geleto et al., 2018), no study has been conducted in Africa’s megacities and those that have been done in urban settings, have either been quantitative studies (Mirkuzie et al., 2014; Nwameme et al., 2014; Worku et al., 2013) or qualitative studies with health care providers (Afari et al., 2014; Austin et al., 2015; Chi et al., 2015). Only two studies recruited the women who actually travelled to the health facilities (Echoka et al., 2014; Ganle et al., 2014), and these studies looked at barriers to access and utilisation of care only, with no focus on how women reached facilities in the broader sense. Neither of these studies attempted to explain the decision-making process of women as it relates to the choices made when travelling to health facilities while in situations of emergency. This paper attempts to fill this gap in the literature. Specifically, our objective was to explore in granular details the second delay of care with a focus on the travel of pregnant women in emergency situations within a megacity, using Lagos, Nigeria as a case study. It is expected that insights from this paper will be relevant for planning service delivery and policy initiatives in similar settings.

### 2. Research context

Lagos, South-West Nigeria, West Africa is bounded to the south by the Atlantic Ocean, north and east by other states in Nigeria, and to the west by Benin Republic. It is divided into 20 local government areas (LGA). In terms of its population, the National Population Commission of Nigeria estimates that 17.5 million reside in Lagos. This figure is however disputed by the state government which puts the population of Lagos at 21 million, with women constituting about half of it (Lagos Bureau of Statistics, 2013; United Nations, 2019). Some researchers have suggested that the population of Lagos will be tripled by 2050 (Hoornweg and Pope, 2017). Since its annexation by British colonialists in 1861, Lagos was always intended to be an urban settlement (Olalekan, 2011). Lagos is the economic nerve centre of Nigeria and arguably the most industrialised part of the country (Chima, 2013). The coastal state has a mix of remote and built-up areas, metropolis and slums, land and riverine areas with a range of travel options including road, water and rail. The most popular mode of travel is by road.

As per the most recent National Demographic and Health Survey, health facility-based delivery in Lagos is 77%, which is in the second highest quintile compared to other parts of the country (National Population Commission and ICF International, 2019). Its estimated maternal mortality ratio (MMR) is 450 (95% CI 360:530) per 100,000 live births (Oye-Adeniran et al., 2011). When disaggregated by LGAs, MMR ranges from 356 per 100,000 live births in Ikeja LGA to 826 per 100,000 live births in Alimosho LGA. A 2014 survey revealed that 306 (1.0%) of 29,988 adult respondents had married sisters who died during pregnancy (45.1%), childbirth (34.9%) or during the six weeks post-childbirth (19.9%) (Odeyemi et al., 2014). Similar to global patterns, hypertension, spontaneous abortions and ectopic pregnancies were the most commonly reported causes of death during pregnancy, while haemorrhage and prolonged or obstructed labour were more commonly reported as causes of death during childbirth in Lagos (Odeyemi et al., 2014; Okonofua et al., 2017).

In terms of available CEmOC facilities, there are 22 secondary health care facilities and one tertiary hospital owned by the state government, as well as one secondary health facility and one tertiary facility owned by the federal government, which are expected to provide EmOC services 24 hours a day [Figure 1]. There is also a complement of three military hospitals and about 35 private hospitals that can be classified as CEmOC facilities with consultants who can provide required EmOC services 24 hours a day, as per State Ministry of Health database. For this study, we have focused only on public sector hospitals that provide CEmOC, as they form the bedrock of universal health coverage in LMICs (Sachs, 2012).

**Figure 1:**
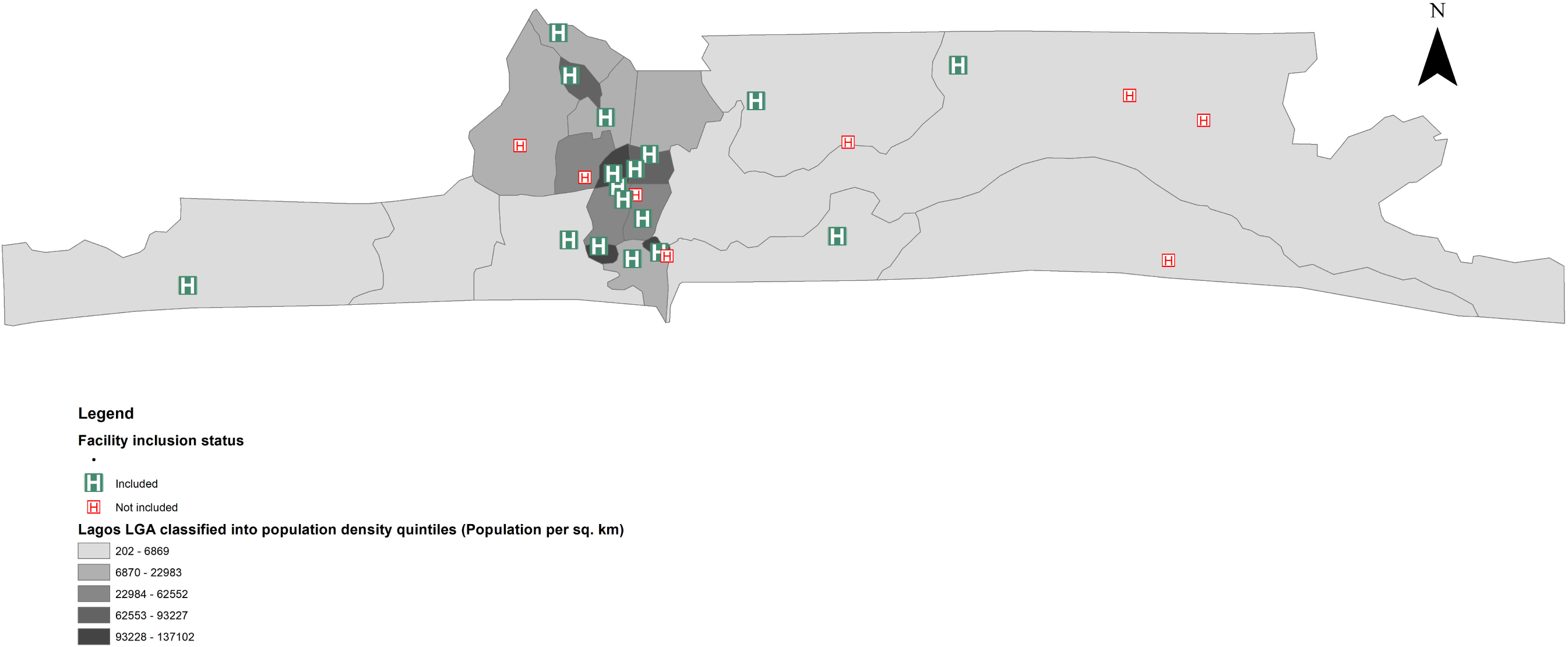
Public sector EmOC facilities in Lagos highlighting those included in this study

## 3. Methods

### 3.1 Sampling and data collection

This study is the qualitative research component of a larger mixed methods study that explored the issues around geographical accessibility to EmOC in Lagos. All government-owned (state and federal) health facilities with capacity to provide the full gamut of CEmOC in Lagos were eligible to participate in this study. Using criterion sampling (Patton, 2015), sixteen of all eligible facilities were purposefully selected for this study. Criteria upon which facilities were sampled included type of urban settlement where the facility is located (town/suburb/city) and type of urban residential area served (slum/non-slum). This was done to ensure maximum variation of travel scenarios of women to facilities. Data was collected between September 2019 and January 2020.

As we could not be physically present in selected facilities at all times, the study involved collaboration with resident doctors and senior medical officers working in obstetrics and gynaecology units and who are typically on duty when pregnant women in situations of emergency present at the health facilities. We requested them to inform the research team when any woman of potential interest presented in the facility. Women and their relatives, 18 years or older, were purposively and opportunistically sampled while ensuring heterogeneity of interviewees in order to guaranty variation of characteristics based on age, presenting complaint, parity and socio-economic status (SES). We borrowed insights from studies conducted in Nigeria to classify women into low, medium or high SES, considering their level of education, employment status and family monthly income (Chukwuonye et al., 2013; Oseni and Odewale, 2017; Oyedeji, 1985). In all, our sample’s variability allowed for an examination of how these characteristics influence travel in situations of emergency. Across all facilities, we interviewed 47 women and 11 relatives of these women who presented at CEmOC facilities in situations of emergency and had received at least one of the nine WHO recommended EmOC signal functions. In every facility, between two and four women were sampled. Table 1 shows the sociodemographic and obstetric profile of the interviewees.

**Table 1:**
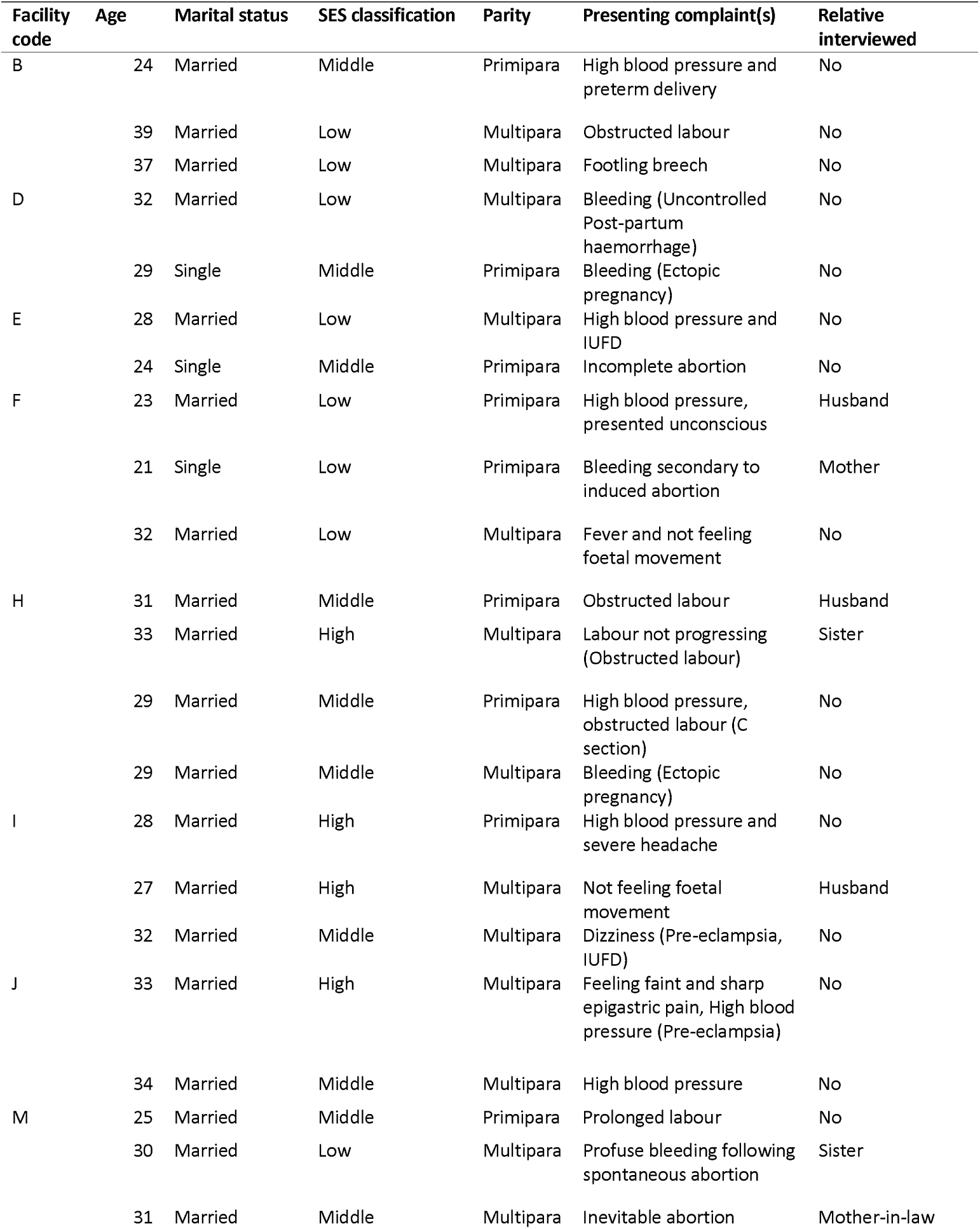
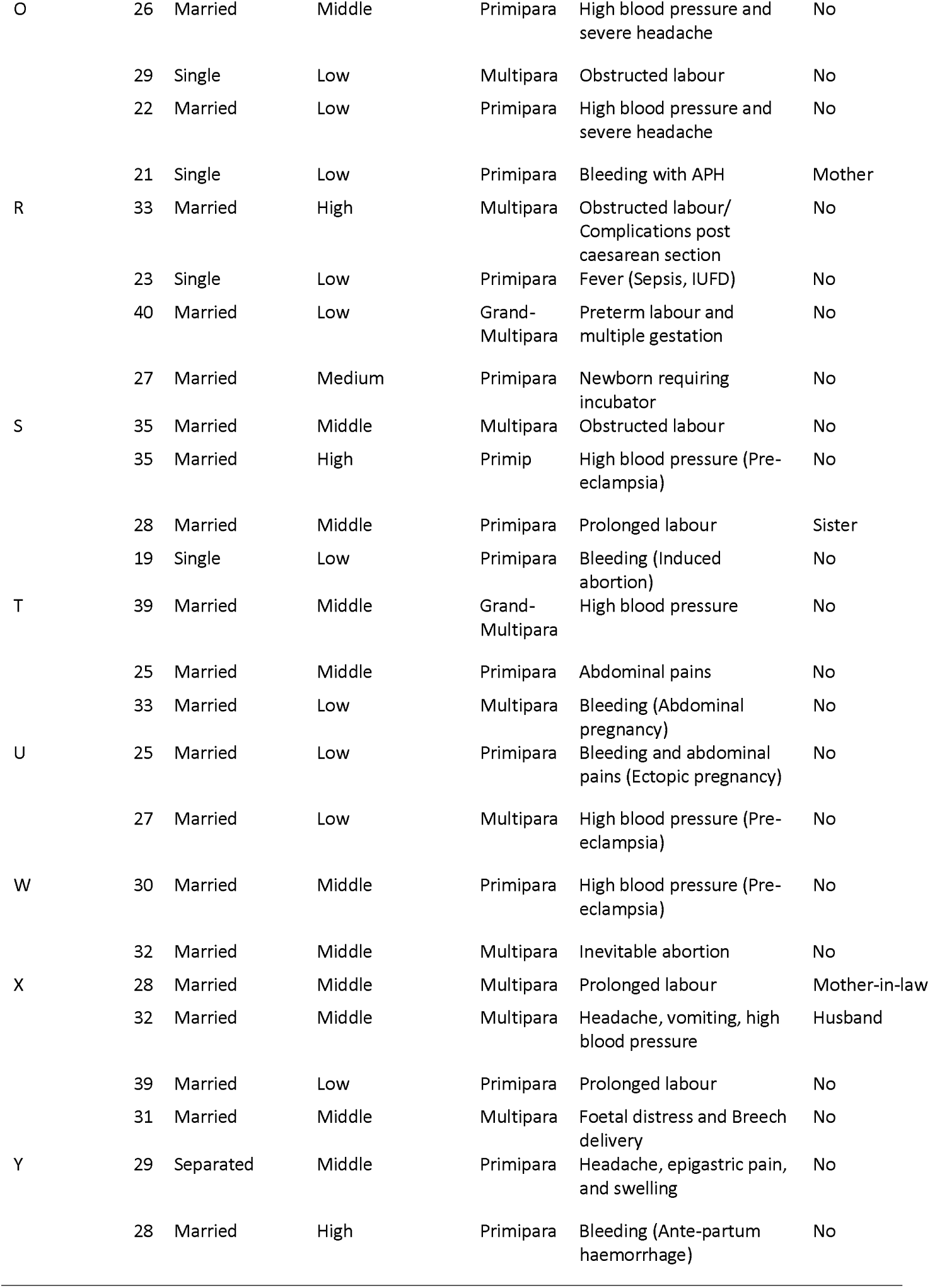
Sociodemographic and obstetric profile of interviewees

Women were approached in the post-natal or recovery wards following permission of the medical team managing their care. Keeping in mind the “baby blues syndrome”, which suggests that 50%– 80% of women have mood swings by the third day postpartum (Fossey et al., 1997; Martinez et al., 2001), we approached only women who had spent more than a day but less than three days on the postnatal ward for this study.

Five interviewers, trained in qualitative research methods and able to communicate effectively in English and the local languages principally spoken in Lagos (*Yoruba* and *Pidgin English*), conducted the in-depth interviews (IDI). We employed IDIs for data collection as they are ideal for preserving confidentiality, especially for a sensitive and emotive issue as the one explored in our research and also allowed a robust exploration of each woman’s experience (McGrath et al., 2019).

Semi-structured topic guides developed first in English language, then translated to *Yoruba* and *Pidgin English*, and subsequently back translated, and tested with non-recruited respondents were used to collect information from the women. Specifically, the topic guide explored issues around planning for travel in situations of emergency, decision-making in such situations, experiences of travel on the day of emergency, challenges and how they were dealt with as well as perceived impact of travel on pregnancy outcomes. Concepts and experiences that we identified in earlier interviews were explored further with participants we interviewed later. Interviews lasted between 20 and just over 40 minutes and were conducted in private rooms of the wards, or in secluded bed areas of the women ensuring that conversations were discrete and private. All interviews were audio-recorded with respondent’s permission using two different dictaphones to provide back up in case of any malfunction and reflective notes capturing subliminal non–verbal events were taken. Only the researchers and participants were present during the sessions. Data collection continued till thematic saturation was achieved.

### 3.2 Ethical considerations

Informed consent was obtained from respondents and no financial incentives offered. Respondents were only recruited if they voluntarily decided to partake in the research. They had the opportunity to withdraw from the study at any time, if they were not able to continue the interview. Those who agreed to partake but were deemed emotionally fragile post-delivery were excluded and offered professional psychological support and counselling. In addition, anonymity of patients and facilities from which they were recruited was maintained in reporting this study.

We obtained ethical approval for the study from the Research and Ethics Committee of the Lagos State University Teaching Hospital (LREC/06/10/1226) and Health Research and Ethics Committee of the Lagos University Teaching Hospital (ADM/DCST/HREC/APP/2880). Social approval for the study was received from the Lagos State Government (LSHSC/2222/VOLII/107).

### 3.3 Data analysis

Following verbatim transcription of audio-recordings, we followed Braun and Clarke’s six-steps for thematic analysis (Braun and Clarke, 2006). To ensure analytical rigour, each transcript was read over by the researcher who conducted the interview and the lead author (ABT), preliminary thoughts on the transcript were collected during a debrief session after interviews had been completed in each facility. These sessions provided an opportunity to check understanding, reflect on emerging themes, identify novel lines of enquiry, and interrogate any peculiarities in the data.

Initial set of codes were generated and applied to the transcripts on subsequent readings. An inductive analytical approach was taken in generating the codes facilitated by a Computer Assisted Qualitative Data Analysis Software, NVivo 11 Plus (QSR International, Memphis, USA). Codes that shared similar meanings were grouped together, and these groupings were reviewed and revised until the coded data had been organised into a set of internally consistent themes. Emerging themes were reviewed in tandem with transcripts to check that they accurately captured content, new understanding tested and alternative explanations sought (Marshall and Rossman, 1999). In describing the emerging themes, constant comparison and deviant case analysis techniques were applied, exploring any variations based on the varying criteria used in data collection. To achieve this, we leveraged our initial categorisation of sampling of included facilities and women to explore similarities and differences between interviewees. In reporting, we followed guidelines from the consolidated criteria for reporting qualitative research (COREQ) checklist (Tong et al., 2007).

## 4. Results

### Decision-making in emergency situations

Pregnant women have several decisions to make when faced with situations of emergency. First, they have to decide when to travel, where to travel and how to reach EmOC providing health facilities. Women in our study did not typically make travel plans for emergencies that could occur in pregnancy before they found themselves in situations of emergency, irrespective of their ante-natal clinic (ANC) attendance. Most of the reported lack of preparation was founded on religious beliefs. One 30-year-old, middle SES, primiparous woman, who presented with increased blood pressure at Facility W (suburb, slum) said, *“Planning [for emergency] means I am expecting a problem. I know that women can have emergencies in pregnancy, but I know my own cannot be like that. God knows why this one happened”*. However, irrespective of age, SES, or parity, women generally acknowledged that they recognised symptoms such as sharp abdominal pain, bleeding, severe headache, fever, not feeling foetal movements, or appearance of foetal parts from the introitus as symptoms that meant they needed to get help. While those who had attended ANC in some health facility (public or private) appeared to have a better sense of urgency, those who had not registered in a hospital before they found themselves in situations of emergency (i.e. un-booked) appeared slower in making the decision to travel to a facility. One relative of an un-booked woman said:

> *“On her arrival, first day, second day, she told me to give her a cloth. I asked why and she said she was seeing something that looked like her menses coming from her vagina… On the third day, we were waiting by the roadside for a vehicle to take our younger one back to where he came from. She just suddenly saw blood and then she alerted me. So, I said she should go inside the house into the bathroom while I go get some water for her. Before I got back to the bathroom, the paint bucket that she sat on was filled with blood. Then she said she was starting to feel dizzy. So, I told her to allow me quickly purchase milk for her. Before I got back from where I went to get the milk, she had already fainted. I thank God, after we poured water on her, screamed her name and also by the help of God, she regained consciousness (Sighs!) Then I started shouting, “Where is a hospital? Where do I take her to”? Then the people described the location of this hospital”. Relative (Sister) of 30-year-old woman, low SES, multipara, spontaneous abortion that had been managed by a traditional birth attendant out-of-state and now presented in the hospital with profuse bleeding [Facility M (suburb, slum)]*.

However, if the emergency was late at night, or there were concerns about security and safety on the roads, some women who recognised that they were in an emergency situation needing facility-based care waited till daybreak to commence their journeys. Women who reported waiting till morning were typically those in early pregnancy (first trimester). However, despite safety concerns, women in later stages of pregnancy (second and third trimesters) reported being more focused on the need to be delivered of the baby than being concerned about security.

> *“I was at home and I was having severe pains from about 2 a.m. that morning. I woke my husband up immediately, but because of our concerns with safety at that time, we decided to leave for the hospital later at about 6 a.m.…”* 29-year-old, medium SES, multipara, with bleeding and abdominal pain [Facility H (suburb, non-slum)]

> *“My husband was saying it is too late and it would not be safe for us to be on the road. But as a pregnant woman, when you are in labour, you are not thinking about anything. You just want them to remove the thing [the baby]”*. 33-year-old, high SES, multipara, labour not progressing [Facility H (suburb, non-slum)]

The choice on when to commence travel did not appear to be influenced by which facility they intended to visit or where it was located. However, there were a few women who knew that they were in an emergency situation but decided to make stopovers along the way to pick up “experienced”, “needed” or “helpful” relatives like mothers and mothers-in-law or go for shopping for items needed for their hospital admission. The choice to do this did not appear to be dependent on SES or parity. A 28-year-old, medium SES, multipara, who presented with prolonged labour said, *“we had to go pick my mother-in-law at [her place] before we started coming to this place [hospital]”. Similarly, another 33-year-old, low SES, multipara, presenting with heavy bleeding [Facility T (suburb, non-slum)] said, “Before I came, I went to market to buy a few things that I thought I needed [in the hospital], before coming here [the hospital]. When I got here, they said they can never let me go, that my BP [blood pressure] was so high at 250 plus [systolic pressure]”*.

For the decision on where to go in situations of emergency, women reported that several factors came into consideration. For some women, the decision on where to go in situations of emergency was made a priori based on the CEmOC facility where they received antenatal care (ANC). For such women, their choice of facility was principally based on proximity to their place of abode as well as community or a relative’s perception of the facility. One 29-year-old, medium SES, primipara, who presented with bleeding as a result of an ectopic pregnancy [Facility Y (suburb, slum)] noted that, *“My sister had a baby here and she recommended it… So, I did not mind registering here. Secondly, this place is near to my house, so just in case something happened, I can quickly get here. And you see, I needed it”*. However, for many women in our sample, perception trumped proximity. For such women, the negative perceptions that they had of specific facilities prevented them from attempting to travel to nearer facilities in situations of emergency. A 24-year-old, middle SES, primiparous who presented with high blood pressure in pregnancy [Facility B (suburb, slum)] said, *“[Facility D (suburb, slum)] was closer to me, but I heard that they exchange people’s babies when they are born… So, me I said, I am not going there”*. A positive perception made women in situations of emergency travel even further out because another facility was deemed to *“be better”, “provide good care”, or as having “caring staff”*. The case for going to a farther hospital was reinforced when influential relatives (husbands and mothers-in-law) or associates (leaders in religious organisations) recommend those facilities. One 25-year-old, low SES, primiparous, bleeding and abdominal pains [Facility U (suburb, slum)] said, *“It was my mother-in-law that suggested that we use this hospital. She told me that is the hospital she uses, and other relatives also deliver here. And that I will receive good care”*. When probed further, we found that there were three closer facilities that were alternatives to her. Another woman said:

> *“It was my husband and brother that brought me here… we passed some [three] hospitals before we got here, but everyone knows that the people here are very caring”. 33-year-old, high SES, multipara, feeling faint and sharp upper abdominal pain [Facility J (suburb, non-slum)]*

Health insurance coverage in specific facilities was an additional reason given by women in deciding where to go. One 32-year-old, medium SES, multiparous, who presented with dizziness and was managed for pre-eclampsia, lost her baby while going to a more distant facility where her health insurance would be accepted. In addition, women talked about having connections (i.e. privileged access) to senior health workers in the facility as a deciding factor. In fact, pregnant women in situations of emergency reported altering their initial choice of where to go while in transit based on advice received from associates and influential people that they know and call for advice in such situations. The underlying motivation appeared to be the reassurance that they would receive the emergency care that they needed urgently upon their arrival at the facility. Use of connections was particularly reported amongst high and medium SES women. In a respondent’s words:

> *“My husband and I agreed that I should do my antenatal at [Facility E (suburb, non-slum)]. But when my husband called his boss that night, he advised that we are better going to [Facility I]. He knew the oga [i.e. medical director] there, and he will call them, so that we get the necessary care.” 27-year-old, high SES, multipara, not feeling baby movement [Facility I (suburb, slum)]*

Women in our study did not report their ‘how to travel’ i.e. mode of transportation as a matter of choice; it was a given based on availability. For those who owned their personal cars, this was the option used in all instances. Some who did not have their own cars relied on partners to first make a trip to loan a car from relatives or friends or to hail a taxi through mobile apps, direct calls to known taxi drivers or through roadside taxis. However, for those who could not access any of the aforementioned means, the next two popular options were public “kękę” (tricycle) and “okada” (motorcycle), with general preference for the former. A 26-year-old, low SES, primipara, who presented with high blood pressure and severe headache [Facility O] said, *“I prefer kękę, it is safer than okada. You know it is emergency. So, with kękę, at least there is cover, because someone can still fall from okada, while dizzy”*.

However, for some who live in riverine areas, the sole mode of transportation was by boat - either one that they owned or one that was used by the public. Nevertheless, this mode of transport still needed to be combined with land travel to reach facilities in situations of emergency.

> *“The only way to come here was to first take boat to the other side and then take kękę from there. We don’t have hospital on our side. My husband has his own boat that he uses for fishing. So, with the help of his brother, they brought me to this hospital…”. 32-year-old, low SES, multipara, with fever and not feeling foetal movement [Facility F (town, non-slum)]*.

### Travel experiences of pregnant women in situations of emergency

Women and their relatives reported varying experiences when they set out to CEmOC facilities. Experiences varied with the time of travel, as well as the geographical terrain navigated to reach the facility. Irrespective of whether the facility was located in a town, suburb or city, women who lived within one or two streets away from CEmOC facilities had about 5-10 minutes of travel using taxis or their personal cars. Across the entire sample of 47 women, estimated time of travel ranged from five minutes to almost four hours in daytime and between five and 40 minutes at night. At night, there did not appear to be much variation in travel time to reach facilities based in towns, suburbs or cities, irrespective of how close or far women lived from the facilities. Neither was there much variation in reported travel time to reach facilities serving mostly non-slum (5-30 minutes) and slum (15-40 minutes) populations. Women reported that time of travel in the afternoon could be between two and six times more if there was heavy traffic. In such conditions, women reported resorting to the use of extreme measures, including driving on illegal lanes, facing oncoming vehicles or abandoning the cars they were in to flag down a motorcycle. One particular woman’s experience trying to reach a facility located in a suburb, non-slum area captures some of these extremes:

> *“The experience of getting to this place was terrible. I first went to a private hospital. From my house to that hospital, typically one hour no traffic. But we needed to be in hurry, we had to take the BRT [Bus Rapid Transit] lane. A policeman stopped us but the church member who was carrying me explained to him that he was carrying a pregnant woman, he then let us pass. But the traffic was still too much. We had to turn back on the road to drive ‘one-way’ to reach this hospital.”* 35-year-old, medium SES, multipara, with obstructed labour [Facility S (city, non-slum)].

In other instances, women reported using motorcycles which allowed them “beat the traffic”. However, they highlighted the discomfort and additional pain they had to endure with this mode of travel, compounded by the deplorable conditions of many of the roads. This road condition concern was shared by women irrespective of the time of the day that they travelled, the length of the trip or the location of the facility. One woman who presented at a facility located in a suburb and non-slum area said:

> *“I had to take okada from my house to the PHC, because I knew this would be faster, especially as the road is not tarred and there is usually plenty traffic on it. In all, the journey was about 30 minutes. On my way going, when the bike enters a pothole, I will shout because I was in pain. But I didn’t have a choice! It was from there that they referred me here”* 31-year-old, medium SES, primipara, with prolonged labour [Facility H (suburb, non-slum)]

Another woman who travelled to reach a facility in a suburb, slum area said:

> *“I took taxi*… *I was worried when I was being transported on the roads. There were so many potholes. I was just holding myself. Even though it was just a short journey, maybe like six minutes, from my house to the hospital. But I felt like I was going to die.” 39-year-old, low SES, multiparous, obstructed labour [Facility B (suburb, slum)]*

The deplorable conditions of the roads only further prolonged travel. One 23-year-old, low SES, primiparous woman who presented at a facility located in a non-slum, town with raised blood pressure [Facility F (town, non-slum)] said that, *“Normally that journey will take maybe 20 minutes, because it is a highway, but my husband said it took us over 45 minutes… It is under construction at the moment”*.

Women who travelled at night and presented at some facilities located in towns and suburbs reported other experiences including difficulty in finding transportation options, being stopped by police en route and the facility gate being closed upon arrival and only opened after beckoning to the security persons at the gate.

> *“… This was now after 11 p.m. When the doctor came, he said they could not handle it and then referred me to this hospital. We looked for kękę, we couldn’t find. When we finally found one, we got on the road but there were so many policemen on the road stopping us. Though they let us go when they saw me. We got here at about 1 a.m. the next morning. The gate was locked, but when my husband knocked, they let us enter. Thank God for my life. I could have died. It is the water that poured, the container did not break”*. 37-year-old, low SES, multipara, with a footling breech [Facility B (suburb, slum)]

As expected, the cost of travel varied with mode of travel and distance covered. Travel with personal cars was described by women as being “at no additional cost, except for the petrol purchased”. However, for low SES women, many complained about the expensive cost of travel, for which they had to sometimes “borrow money”, “plead with the driver”, “take okada” (which was deemed cheaper), or “walk some distance to save money”. It was not unusual for women to use multiple transport means to reach facilities. A relative (mother) of a 21-year-old, low SES, primiparous, who presented with bleeding as a result of antepartum haemorrhage [Facility O (town, non-slum)] submitted that, *“We spent N200 (US$0.55) from the house to the bus stop in kękę. Then from the bus stop to the main stop in the area of the hospital on a 30-minute bus ride, N200 (US$0.55) and then okada to the hospital, N100 ($0.27) per person”*. Thirty-three of the 47 women in our study who took public transport reported spending between N300 ($0.82) and N7,500 ($20.60) to reach facilities in Lagos. When disaggregated, women who travelled within towns or suburbs to reach CEmOC facilities in cities using public transport appeared to spend more compared to those who travelled within the cities. Twenty-seven of the women who reside in suburbs spent between N1,200 ($3.30) and N7500 ($20.60) while six of their counterparts who reside in urban areas spent between N300 ($0.82) and N2,500 ($6.87). Irrespective of settlement type, women reported that drivers hiked fares if they were carrying luggage with them or if they were travelling at night.

> *“We took taxi to [this hospital] when I started having problems…It would have been N2,000 ($5.49) in the morning but at that time of the night they said N5,000 ($13.74). He even said he could have charged us more but because he is of the same ethnicity as my husband, he allowed us pay less”*. 32-year-old, low SES, multiparous, with bleeding [Facility D (suburb, non-slum)]

Women highlighted that having relatives with them along the way to reach CEmOC facilities was “very helpful”, “important” “most needed”. Relatives reported that they had to try all they could to support and manage their loved ones in situations of emergency and in some cases, they had to treat them any way that they knew. A relative (husband) of a 23-year-old, low SES, primiparous, who presented unconscious at Facility F (town, non-slum) said, *“She definitely does not know how she got here. She was gone [unconscious]! I had to put a spoon in her mouth [a local technique done to unconscious patients to prevent the teeth from apposing which signifies end of life]”*.

### Reaching another facility after being referred

Seventeen women in our study required referral to a CEmOC facilities when they presented with emergencies, as the basic or comprehensive EmOC facility they initially presented to could not provide the necessary care. Service delivery limitations included lack of bed space or incubators for their newborns. Some of these women had to move through multiple facilities in order to access the required care. A 24-year-old, medium SES, primipara, who presented with high blood pressure and pre-term delivery in Facility B (suburb, slum) stated *“I registered with one matron in my community and I was placed on blood pressure medications. But the baby was still small and needed incubator. So, she referred me to [Facility D (suburb, non-slum)] … But when I got to that facility, the doctor said, they don’t have any space for incubator, so they now referred me here [Facility B]”*.

Ambulances, which are expected to be available in all state-owned CEmOC facilities, are typically used to transport patients from facility to facility in situations of emergency. When available and not in use and if there was no traffic, women referred in hospital ambulances found it to be advantageous, irrespective of urban settlement type. The reason given by women for such perception was that an ambulance provides supportive equipment and hospital drivers are more conversant with the directions to the referral hospital. However, some women reported that even though they had to pay to be transported in the hospital ambulance, it did not help to reach the referral facility any quicker. The women attributed this to the lack of regard for ambulances by other road users; there does not appear to be a culture of motorists giving way to ambulances on Lagos roads.

> *“The C-section was done at [Facility X (suburb, non-slum)] and then afterwards I noticed that I was coughing blood and they had to transfer me to this hospital [Facility R (city, non-slum)]. My husband, one nurse and a doctor were with me in the ambulance. The road was free… maybe 15 minutes max because the road was clear. It (the ambulance) had all I needed – oxygen, life support, everything! And then I have never been here in my life, so not like I even know my way to this place. It was a smooth process!”* 33-year-old, high SES, multiparous, with complications post-caesarean [Facility R (city, non-slum)]

> *“They used their ambulance, which they told me to pay for. I paid N3,000 ($8.25). I left the hospital at 4 p.m. and I got here around 9 p.m. The traffic was really bad that day!! The ambulance did not help us to get here faster. Even people in their private car were dragging the road with us. They don’t even want to know if somebody is dying or something… If it was in my state, people will give way to ambulance”*. 24-year-old, single, primipara, with incomplete abortion [Facility E (suburb, non-slum)]

Some women were also referred to state-owned CEmOC facilities from primary health care facilities or private hospitals. However, ambulance use for referral in these facilities was variable. In many instances, women were referred and expected to figure out on their own how to reach the referral facility. One 27-year-old, medium SES, primiparous, who was referred from a health centre with her newborn requiring incubator [Facility R (city, non-slum)] said, *“They did not even mention ambulance. So, my husband had to get taxi. We travelled for almost three hours from [the health centre] to this hospital”*.

## 5. Discussion

This study set out to critically explore the factors influencing and experiences of travel of pregnant women in situations where they are attempting to access EmOC in Africa’s largest megacity, Lagos. Findings showed that after pregnant women have identified that they are in an emergency situation, they are still faced with real conundrums on “when”, “where” and “how” to travel to reach facilities that are able to provide the care they need. While this choice-making is a shared activity amongst all women, the choice-options and the process of making the choice vary. However, our interviews revealed remarkably similar experiences of travel, irrespective of women’s age, socio-economic background, or obstetric history. Nevertheless, the extent to which travel experiences impacted their ability to reach EmOC facilities depended more on individual agency and support systems. Due to service delivery limitations, some women had to be further referred from one facility to another, which further prolongs travel and sometimes increases cost.

Contrary to suggestions in the literature that uneducated women tend to delay in recognising danger signs (Mutua et al., 2015; Oiyemhonlan et al., 2013), women in our study recognised danger signs, regardless of their age, SES, obstetric history and presenting complaint. However, our findings showed that being aware of danger signs does not necessarily translate to responding to the urgency of the situation. It is difficult to conclude that decision on when to travel was made in quicker time amongst those of higher SES compared to low SES in our study. Our finding points to the reluctance of women facing obstetric emergencies, many of whom are classed low SES, in presenting at facilities, when they had not registered for ANC. This might be due to concerns at being chastised for not being booked or the cost associated with facility-based care (Wright et al., 2017). On the other hand, some of those who were classed as high SES also delayed in deciding when to travel either because they felt it was best to travel when it was safer (i.e. during the day) or because they downplayed indications of their being in danger. Women who lost their lives due to complications of pregnancy and childbirth in Gambia have been shown to underestimate the severity of their complication and as such delay travel and care seeking (Cham et al., 2005). Our findings indicate that the perception of urgency or severity may be influenced by some symptoms which women perceive as being more ominous than others. For example, excessive bleeding is clearly considered a danger as against being told that one has high blood pressure when not feeling any specific symptoms.

The choices that women made on ‘where’ to go was influenced by proximity for some women, especially when they resided close to facilities. However, for many women, proximity was trumped by family or community perception of the facility, health insurance coverage, connections with highly placed individuals within the health system and the advice of influential people. While a previous study showed that there is a general “positive perception” by women of public CEmOC facilities in Lagos, especially as it relates to the availability of technically sound skilled health personnel (Wright et al., 2017), concerns remain regarding responsiveness of staff and cost of service utilisation. Evidence points to the consensus that women in LMICs place importance on attitudes of health care providers (HCPs) (Bohren et al., 2015; Okewole et al., 2013). Women in our study reported travelling even farther to CEmOC facilities for these reasons. Additionally, some respondents reported changing their course of travel because of links with influential people at specific hospitals. While a recent systematic review on barriers to EmOC access in sub-Saharan Africa does not point to lack of connections as a barrier (Geleto et al., 2018), women, especially high SES ones, clearly felt that they needed the “added advantage” it provides, as a way of guaranteeing the quality of care they would receive.

Motorised and non-motorised options have been reported as options of travel available to pregnant women in emergency situations living in LMICs (Wilson et al., 2013). In our study, we found that choice-making on “how” to travel was simply based on availability. For the high and medium SES women who owned a car or had a relative nearby who owned a car, or had access to and could afford taxis, vehicles were the choice model of transport in situations of emergency. In other LMIC settings, lack of vehicles has been reported as a barrier to accessing EmOC (Combs Thorsen et al., 2012; Echoka et al., 2014; Ganle et al., 2014; Nwameme et al., 2014; Worku et al., 2013). However, the striking finding in our study was the risk that women in our study were willing and having to take when they did not have a vehicle to take them to facilities. The motorcycle was a common mode of travel for women, especially those in remote areas as well as areas prone to significant traffic. However, it is important to bear in mind that beyond the risk posed by these two-wheelers to pregnant women, at least 30% of road traffic accidents have been attributed to them (Ibrahim et al., 2017). It was therefore good to find that women in our study perceived the tricycle as a safer alternative. This probably complements ongoing efforts to scale up tricycles for supporting pregnant women in emergency in some other parts of the country (Medicins Sans Frontières, 2018).

Irrespective of the choices that women made, their socio-economic status or obstetric history, many women in our study reported facing significant challenges in travelling to health facilities, with different challenges faced during the daytime (traffic congestion) and night-time (security concerns and scarcity of public transportation). The deplorable conditions of many of the roads was a huge challenge, regardless of the time of day. A 2015 review reported travel time ranges between 10 minutes and a day (Wilson et al., 2013). Women in our study reported they spent between 5 minutes and 4 hours. This is probably because of the urban nature of our study setting. The women also reported that the traffic combined with the bad roads could increase their travel time as much as 200-600%. Their estimates are not too far from those reported in the grey literature, where as much as 800% increase in travel time due to traffic has been reported (Akinwotu, 2015). Commuters in Lagos spend an average of 30 hours in traffic every week (Obi, 2018). In a recent study conducted in Nairobi, Kenya, traffic contributed to a 166% and 138% increase in time of travel for patients to reach secondary and tertiary hospitals respectively (Fraser et al., 2020). For the urban poor in particular, such increases in travel time have been found to be a strong deterrent to seeking EmOC. A study in Bangladesh estimated that for every 5-minute increase in travel time to the nearest EmOC facility there is a 30% decrease in the likelihood of delivery at an EmOC facility; favouring home-based care (Panciera et al., 2016). This ultimately nullifies any drive to scale up facility-based deliveries in the absence of broader systemic and infrastructural interventions to address the second delay. In our previously published multi-stakeholder analysis on EmOC access, while the Lagos State government believed that EmOC facilities have been strategically located across the state, several women reported difficulty in accessing facilities (Banke-Thomas et al., 2017). Some have suggested that these constraints could be related to poorly located EmOC services (Niyitegeka et al., 2017) or insufficient numbers of EmOC facilities for the users within a recommended distance (Mkoka et al., 2014).

For night travel, security concerns reported by women is shared by the Lagos population. However, no woman in our study specifically reported this as part of her experience in trying to reach a health facility. The price hike at night is not a unique experience in Lagos. Pregnant women in Ethiopia and Nepal have previously reported feeling financially exploited by transporters (Wilson et al., 2013). Such exploitation was an issue of concern for women who already found day fare rates as prohibitive. Several women in sub-Saharan Africa report cost of travel and lack of transport funds as a barrier to EmOC access (Geleto et al., 2018). However, one key facilitator that in some manner helped to improve experience of travel was having a relative come along on the journey to the facility. Relatives play a significant role providing the emotional, financial and logistical support that women need and for which the health system is not fully structured to provide. A recent qualitative study in Ghana highlighted that sometimes relatives have to play the role of “escort” since health facilities do not have enough staff to escort patients (Daniels and Abuosi, 2020).

As had been previously reported in the literature (Cham et al., 2005), our study also found that even when women made it to a CEmOC facility, referral between facilities further prolonged delays and increased cost of travel for women. Specifically, some women in our study who arrived at secondary facilities were referred to other secondary or tertiary facilities mostly due to lack of bed space or lack of incubators. These referrals from secondary to secondary or tertiary facilities appear to be functional and many pregnant women who were referred and transferred with the available ambulance described the process as an efficient process. However, referrals from primary and private facilities to secondary or tertiary facilities remain fraught with many challenges including lack of ambulances, with women left alone to figure out how they would reach the facilities. Such shortages have been shown to discourage healthcare providers from referring clients (Afari et al., 2014; Ueno et al., 2015). Poor referral systems are huge barriers to accessing EmOC (Chi et al., 2015). The Lagos State Ambulance system mostly functions at the secondary to secondary level, but not so much for referrals from primary to secondary facilities. However, even when an ambulance service is available, it does not always guarantee quicker transit time. This was mostly attributed to other road users not giving way to ambulances. This issue of driver etiquette has long been raised as one that needs addressing in Lagos in an audit of ambulance effectiveness (Adewole et al., 2012) and was also recently reported to be an issue affecting the EmOC referral system in Ghana (Daniels and Abuosi, 2020).

There are clear implications for policy and practice based on our findings, which support the need for governments to target both health systems and the overall SES of women to improve EmOC access (Anto-Ocrah et al., 2020; Kyei-Nimakoh et al., 2017). Firstly, it is important that the practice of birth preparedness and complications readiness (BPCR) as part of routine ante-natal care in Lagos hospitals (Oni et al., 2016) need to be sustained. However, in addition to the danger sign recognition focus of BPCR (“when”), more emphasis needs to be placed on advising pregnant women on “where” and “how” they plan to travel in situations of obstetric emergency (Aduloju et al., 2017). Clearly, there will remain some women who will be yet to commence ANC before they have an emergency, for example, as seen in our study, women with ectopic pregnancies. Sending messages through opinion leaders in the community may be helpful to reach these women. These women can also be reached via mobile phones as was done in Western Kenya (Onono et al., 2019).

Secondly, there is a need to standardise costs to women for receiving EmOC, expand health insurance coverage and fully implement respectful maternity care (World Health Organization, 2018) in public facilities, so that women are not conflicted in their choice-making when in emergency situations. Thirdly, efforts need to be put in place to improve the travel experiences of women while minimising the risk that they may be forced to undertake in situations of emergency. While broad sweep road infrastructural improvements will be helpful to all aspects of state development, this will probably be cost-intensive. Leveraging existing structures, such as establishing partnerships with specific taxi companies and tricycles, might offer a cost-effective and quick gains. However, a recent partial ban of tricycles in Lagos (Olasukanmi, 2020) may mandate some policy reflections, before this can be considered. Indeed, tricycle riders and private taxi drivers can be trained on proper transfer of women in situations of emergency and integrated into the referral process. In addition, indemnity cover to ensure that liabilities that they may incur while transporting women in situations of emergency such as physical damages to their vehicle (Afari et al., 2014), should be covered. Legal permission to women in emergencies to use bus-only lanes when in actual emergency situations can also help reduce travel time in emergencies. Campaign for attitudinal change of drivers, as it relates to giving way for ambulances will also be a helpful measure. However, this needs to be supported by legislation to ban misuse of such rights by ambulance drivers, ensuring that the siren is only used in situations of emergency.

To our knowledge, this is the first qualitative study that rigorously explores issues around travel of pregnant women in situations of emergency in a LMIC megacity. In it, various characteristics of the interviewees were identified including age, obstetric history, presenting complaint, and complication for which they were managed. In addition, facilities in different sites, topographies, and with different population densities were also included. By seeking to maximise the heterogeneity of the sampled facilities and speaking to different women with varying characteristics within each facility, the study reflects multiple experiences of travel and the effect that the various characteristics could have on travel to reach facilities when in emergency. Nevertheless, this study also has limitations. For example, we only focused on women who made it to the facility, excluding those who had emergency experience but never made it to the facility. Future studies need to engage with women and their relatives within the community, in order to identify those who fall in this category and attempt to capture their experiences.

## 6. Conclusion

Our study revealed that the delay in travel to health facilities is a real experience of pregnant women in situations of emergency even in a megacity like Lagos. For pregnant women, reaching a facility in situations of emergency is usually a matter of life and death. However, the ability to reach such facilities should not be based on chance, connections, or capacity. Every woman in such situation needs to have a fair opportunity to reach a facility and receive the necessary care at such critical moments. If the goal remains to leave no one behind, then in addition to other health system strengthening interventions, referral systems need to be improved, advocacy to encourage women to use their nearest facilities when in situations of emergency and partnerships with private sector need to be explored.

## Data Availability

Data will be made available on reasonable request.

